# Association of factors linked to health inequalities and the risk of antibiotic-resistant infection in high-income countries: a systematic scoping review

**DOI:** 10.1101/2025.05.17.25327823

**Authors:** Amrita Ghataure, Ellie L. Gilham, Ella Casale, Eleanor J Harvey, Caroline DeBrún, Viviana Finistrella, Diane Ashiru-Oredope

**Affiliations:** Health Protection Research Unit for Antimicrobial resistant infections and Healthcare-Associated Infection (HCAI), Imperial Colleague London, London, UK; Antimicrobial Resistance and Healthcare-associated Infection Division, UK Health Security Agency (UKHSA), London, UK; Knowledge and Evidence Services, UK Health Security Agency (UKHSA), London, UK; School of Health and Life Sciences, Teesside University, Middlesbrough UK.UK

## Abstract

**Introduction:** Antibiotic-resistant infections disproportionately affect vulnerable populations, yet research on health inequalities in this context remains limited. This scoping review explores the relationship between health inequalities and the risk of antibiotic-resistant infections in high-income countries.

**Method:** A comprehensive search was performed on Ovid Embase, Ovid Medline (R) All, and Google Scholar, including studies published between January 2010 and February 2024 from high-income countries. The search focused on key pathogens identified by the 2018-24 UK AMR National Action Plan. Health inequalities were defined by factors such as socioeconomic status (income, employment), protected characteristics (age, gender, ethnicity), and vulnerable groups (e.g., migrants, homeless individuals, and sex workers). Studies on sexually transmitted, foodborne, viral, or fungal infections, and those from low-or middle-income countries were excluded.

**Results:** Of 203 papers reviewed, 18 were included. Most studies were from the U.S (12), followed by the UK (2), New Zealand (2), Australia (1), and a European study. The most frequently studied pathogen was Staphylococcus aureus, with ethnicity, socioeconomic deprivation and age being the primary factors explored. A few studies also considered migration status.

In the U.S., Black patients exhibited higher Methicillin-resistant Staphylococcus aureus (MRSA) infection rates compared to White patients, with rates 2-3 times higher despite an overall decline in MRSA. Indigenous Australians similarly had higher MRSA rates.

Socioeconomic factors, such as deprivation and age, were significant risk factors; however, when accounting for these factors, racial disparities in MRSA were significantly reduced.

For Streptococcus pneumoniae infections, Hispanic patients showed higher penicillin resistance than non-Hispanic White patients. Escherichia coli resistance was more prevalent among lower-income individuals or those in high-deprivation areas, with ethnic minorities in New Zealand also disproportionately affected. Additionally, Helicobacter pylori infections were most common in Māori individuals in New Zealand, and income inequality was strongly linked to antibiotic resistance in Enterococcus, Klebsiella, and Pseudomonas in Europe.

**Conclusion:** This review highlights the potential impact of factors associated with health inequalities on the risk of antibiotic-resistant infections, with higher rates of resistant infection seemingly associated with ethnic minorities, especially Black and Indigenous populations, as well as with income inequalities.

## Introduction

Antimicrobial resistance (AMR) is a significant threat to modern medicine and global health, jeopardising the treatment of both communicable and non-communicable disease.^1^ It is estimated that antibiotic resistance alone is responsible for 1.27 million deaths globally, surpassing the mortality rates for HIV/AIDS, breast cancer or malaria.^2^ Infections caused by resistant bacteria result in increased morbidity and mortality, prolonged hospital stays and increased health costs.^3^ AMR is driven by a complex interplay of proximal and distal factors across biological, social and environmental settings; a universally recognised driver is the inappropriate use of antibiotics in human and animal populations.^4,5,6^ While high-income countries report the highest rates of antibiotic consumption compared to low– and middle-income countries,^7^ reducing antibiotic use alone is insufficient to control AMR, in part due to the continued transmission of resistant strains and genes.^8^

Avoidable health inequalities exist across social groups, affecting disease prevalence, overall health, and access to quality healthcare.^9,10,11^ These disparities stem from a combination of structural and individual factors—including education, housing, employment, and living conditions—collectively known as the social determinants of health, which are shaped by political, social, and economic forces.^12^

Individuals in lower socioeconomic positions face an increased risk of poor health, encompassing both chronic and infectious diseases. However, the relationship between health inequalities and AMR is complicated and multifactorial. Evidence demonstrates that COVID-19 and other infectious diseases disproportionately affect vulnerable and disadvantaged groups, such as those living in poverty and overcrowded conditions.^13^ A review of 108 systematic reviews in high-income countries revealed a higher risk of infectious diseases, AMR, and lower vaccination uptake among individuals with lower incomes, lower educational attainment, higher area-level deprivation, lower socioeconomic status, and poor living conditions.^14^ While evidence specifically on AMR was limited, it is reasonable to expect similar patterns, as the risk of acquiring resistant infections is intertwined with the social determinants of health. For instance, disadvantaged populations experience differential rates of exposure and susceptibility to infections due to their living conditions, increased psychological stress, and reduced access to vaccination, all of which contribute to disease incidence. Furthermore, variations in antibiotic prescribing patterns exist according to locality and deprivation, with the highest levels of antibiotic prescribing in England occurring in the most deprived areas.^15^ Addressing these disparities is vital for improving health outcomes and mitigating the burden of AMR.

Despite emerging evidence on health disparities in infectious diseases, research specifically examining their impact on AMR remains scarce. This scoping review was one of three identified questions that form part of a wider study initiated in 2019 following the establishment of a health inequality specific workstream within the Healthcare Associated Infection and AMR division of UKHSA.^16,17^ It aims to synthesise existing evidence on the association between socioeconomic status, deprivation, protected characteristics (such as age, sex, disability, race, religion, or sexual orientation), vulnerable group status, and geography with the risk of acquiring an antibiotic-resistant infection. Although low– and middle-income countries bear a higher burden of infectious diseases, this review focuses on high-income countries due to their distinct challenges, including variations in healthcare access, antibiotic prescribing practices, and socioeconomic inequalities. By analysing literature from both hospital and community settings across diverse age groups and key pathogens, this review seeks to identify gaps in knowledge, inform policy, and highlight strategies to address AMR through a health equity lens.

## Methods

This scoping review was informed by the protocol published by Lishman *et al.*^18^ The study followed the Preferred Reporting Items for Systematic Reviews and Meta-Analyses (PRISMA) statement extension for Scoping Reviews (PRISMA-ScR).^19^

### Research question

We sought to answer:

What evidence exists for an association between elements of health inequalities and the risk of antibiotic-resistant infection in high-income countries?

Elements of health inequalities included:

- Socioeconomic status and deprivation (Employment status, income levels, deprivation categories).
- Protected characteristics (Age, Gender, Ethnicity, Sexual orientation, Disability).^20^
- Vulnerable groups (Migration status, sex workers, people who inject drugs (PWIDs), the homeless).
- Geography (Urban or rural dwelling).

### Search strategy

We carried out a scoping review using the terms in Appendix 1. The initial search included literature published between 1 January 2010 to 29 April 2021, with the search updated to include papers published up to 7 February 2024, limited to English-language studies. The search was performed in Ovid Embase, Ovid Medline (R) All, and Google by a Knowledge and Evidence Specialist. A pilot search was reviewed by the project lead to ensure relevance. To maintain focus on antibiotic resistance, the terms *COVID-19* and *pandemic* were excluded.

An initial screening process was conducted to remove irrelevant or duplicate studies. Additionally, a snowballing technique was employed to identify relevant studies from the reference lists of included papers.

### Selection criteria

After an initial screening by a Knowledge and Evidence Specialist, a single reviewer assessed studies against predefined inclusion and exclusion criteria (Table 1).

**Table 1.**
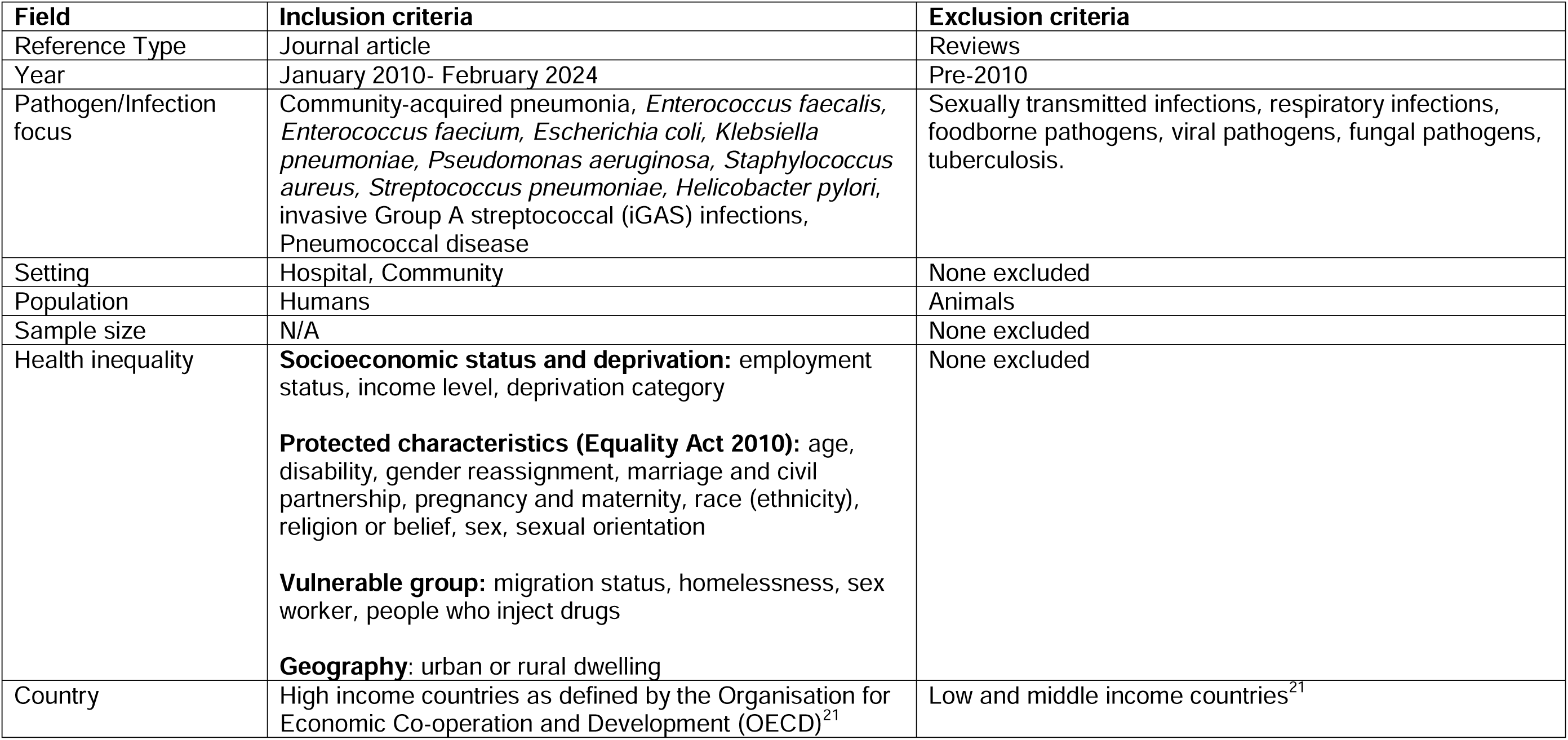
Inclusion and exclusion criteria.

| Field | Inclusion criteria | Exclusion criteria |
| --- | --- | --- |
| Reference Type | Journal article | Reviews |
| Year | January 2010- February 2024 | Pre-2010 |
| Pathogen/Infection focus | Community-acquired pneumonia, *Enterococcus faecalis*, *Enterococcus faecium*, *Escherichia coli*, *Klebsiella pneumoniae*, *Pseudomonas aeruginosa*, *Staphylococcus aureus*, *Streptococcus pneumoniae*, *Helicobacter pylori*, invasive Group A streptococcal (iGAS) infections, Pneumococcal disease | Sexually transmitted infections, respiratory infections, foodborne pathogens, viral pathogens, fungal pathogens, tuberculosis. |
| Setting | Hospital, Community | None excluded |
| Population | Humans | Animals |
| Sample size | N/A | None excluded |
| Health inequality | **Socioeconomic status and deprivation:** employment status, income level, deprivation category<br>**Protected characteristics (Equality Act 2010):** age, disability, gender reassignment, marriage and civil partnership, pregnancy and maternity, race (ethnicity), religion or belief, sex, sexual orientation<br>**Vulnerable group:** migration status, homelessness, sex worker, people who inject drugs<br>**Geography:** urban or rural dwelling | None excluded |
| Country | High income countries as defined by the Organisation for Economic Co-operation and Development (OECD)<sup>21</sup> | Low and middle income countries<sup>21</sup> |

Studies that met exclusion criteria were removed. Pathogens and infections of interest were selected based on the UK Antimicrobial Resistance (AMR) National Action Plan 2019-24, focusing on key organisms relevant to antibiotic resistance.^1^

### Data extraction

A data extraction table was developed using Microsoft Excel and included 17 fields: reference type, author, year of publication, title, journal, pathogen, key findings, setting, population, elements of health inequalities covered, country, UK region, exclude/include, keywords, abstract, URL and database. Results were reviewed at random by the project team at fortnightly meetings to reach a consensus decision.

## Results

The initial search returned 203 records. A first screen excluded 133 records, comprising 118 irrelevant records and 15 duplicates (Figure 1). The remaining 71 results were then screened for relevance to resistant bacterial infections, health inequalities and studies in high-income countries, resulting in 21 articles being screened further for pathogens of interest. Systematic reviews and meta-analyses were extracted for separate analysis (6/71 records). Seven papers were marked as undecided by the reviewer and subjected to a second review. Subsequent screening for the pathogens of interest listed in Table 1 resulted in the inclusion of 10 studies in the review (see Supplementary Table 1). An additional 8 papers were identified from reference lists/snowball technique. In total, 18 papers were included in the review.

**Figure 1.**
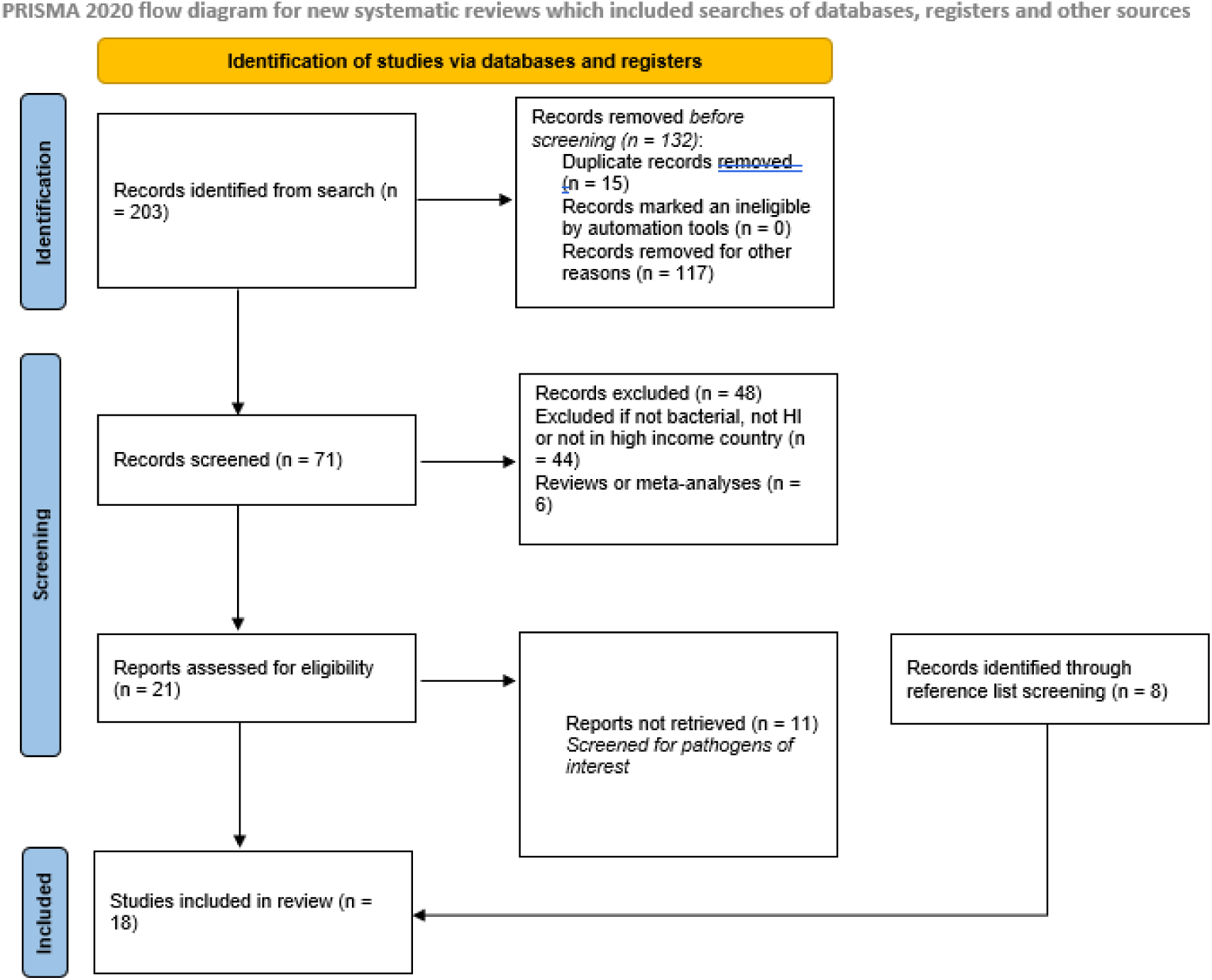
PRISMA flow diagram of studies identified via the literature search.

Study characteristics and findings are outlined in Table 2. Each of the 18 papers were published in a different journal, including the high impact journals PloS One and Clinical infectious disease. Papers were published from 2010 to 2024; with two papers having data collected pre-2010. The majority of studies took place in the United States (12/18) with the remaining covering Europe (multiple countries), Australia, the UK and New Zealand.

**Table 2.** Study characteristics and findings.

| Author | Country | Period assessed | Study design | Target population | Outcome measures | Findings |
| --- | --- | --- | --- | --- | --- | --- |
| ***Staphylococcus aureus*** |  |  |  |  |  |  |
| Ray et al (2012) | USA | 1998-2009 | Retrospective observational study | Insured patients within a large, integrated healthcare system (hospital and community) in Northern California | Percentage of *Staphylococcus aureus* isolates that were MRSA vs. MSSA; incidence of *S. aureus* bloodstream infections | **Race:** African-Americans had a significantly higher likelihood of MRSA infections (OR 1.73, CI 1.64-1.82). Hispanics also showed a higher likelihood (OR 1.11, CI 1.06-1.16). Asians had a lower likelihood of MRSA (OR 0.69, CI 0.65-0.73). Hospital-onset infections were more likely to be MRSA (OR 1.58, CI 1.46-1.70).<br>**Age:** 18-50 years had higher MRSA odds (OR 1.20); 5-18 years had lower odds (OR 0.84), compared to 65+ years.<br>**Deprivation:** Lower income census blocks had higher MRSA odds (OR 1.54 for lowest quintile vs. highest) |
| Gualandi et al. (2018) | USA | 2005-2014 | Retrospective analysis of surveillance data | Residents of selected metropolitan areas within 9 US states, with a focus on Black and White racial groups. | Incidence rates of invasive MRSA infections (hospital-onset, healthcare-associated community-onset, and community-associated) by race; adjusted rate ratios (aRRs) of MRSA in Black patients vs. White patients. | Overall invasive MRSA rates decreased during 2005-2014, but racial disparities persisted.<br>**Hospital-onset (HO) MRSA:** Black patients had significantly higher rates (aRR 3.20, 95% CI 2.35-4.35).<br>**Healthcare-associated community-onset (HACO) MRSA:** Black patients had significantly higher rates (aRR 3.84, 95% CI 2.94-5.01).<br>**Community-associated (CA) MRSA:** Black patients had significantly higher rates (aRR 2.78, 95% CI 2.30-3.37).<br>**Sub-group analysis- Chronic dialysis patients:** Black patients had higher HACO MRSA rates (aRR 1.83, 95% CI 1.72-1.96) |
| Iwamoto et al (2013) | USA | 2005-2010 | Population-based surveillance | Paediatric patients with invasive MRSA infections. | Incidence of invasive MRSA infections (hospital-onset, healthcare-associated community-onset, community-associated) in children. | Incidence of community-associated (CA) MRSA increased (1.1 to 1.7 per 100,000 children). No significant trends in hospital-onset or healthcare-associated community-onset MRSA.<br>**Age:** Infants <90 days had higher incidence (43.9 per 100,000) than older children (2.0 per 100,000).<br>**Ethnicity:** Black children had higher incidence (6.7 per 100,000) than other races (1.6 per 100,000). |
| Tong et al. (2012) | Australia | 2007-2010 | Prospective observational study | Patients with *Staphylococcus aureus* | Incidence of community-associated MRSA (cMRSA) | **Ethnicity:** Age-standardized incidence rate ratio (IRR) for community-associated MRSA (cMRSA) in Indigenous vs. non-Indigenous Australians: 29.2. |
|  |  |  |  | bacteraemia (SAB). | bacteraemia. | Higher proportion of cMRSA in Indigenous patients (24%) vs. other ethnicities (7%). |
| See et al. (2017) | USA | 2009-2011 | Retrospective cohort | Invasive community-associated MRSA cases. | Incidence of community-associated MRSA infections. | **Ethnicity:** Annual invasive community-associated MRSA incidence was higher among Black persons (7.60 per 100,000) than White persons (4.59 per 100,000) (RR, 1.66; 95% CI, 1.52–1.80).<br>**Socioeconomic factors (deprivation)** explained 91% of the racial disparity. Black race was not significantly associated with MRSA incidence after accounting for socioeconomic factors (RR, 1.05; 95% CI, 0.92–1.20). |
| Auguet et al. (2016) | UK | 2011-2012 | Spatial analysis of cross-sectional data | Residents of Lambeth, Southwark, and Lewisham boroughs in South East London | Incidence of HA-MRSA and CA-MRSA | **Deprivation:**<br>-CA-MRSA was associated with household deprivation (RR: 1.72 [1.03–2.94]), household overcrowding (RR: 1.58 [1.04–2.41]), a combined score for overcrowding, low income, and homelessness (RR: 1.76 [1.16–2.70]), and recent immigration to the UK (RR: 1.77 [1.19–2.66]).<br>-HA-MRSA was associated with household deprivation (RR: 1.57 [1.06–2.33]), poor health (RR: 1.10 [1.01–1.19]), and residence in communal care homes (RR: 1.24 [1.12–1.37]).<br>CA-MRSA showed significant spatial clustering, indicating community transmission, while HA-MRSA did not. |
| Jenks et al. (2016) | USA (New York) | 2011-2013 | Cross-sectional observational study | Patients with Skin and Soft Tissue Infections (SSTIs) at 6 New York City-area Community Health Centres (CHCs). | Prevalence of CA-MRSA and MSSA in wound and nasal cultures | **Migrant populations:** Immigrants had lower rates of MRSA wound infections than native-born patients. Immigrants had significantly higher rates of MSSA wound cultures (OR=3.5, 95% CI: 1.3, 9.7). |
| Immergluck et al. (2023) | USA (New York) | 2015-2017 | Cross-sectional observational study | Patients with Skin and Soft Tissue Infections (SSTIs) at 6 New York City-area Community Health Centres (CHCs) and hospital emergency departments. | Prevalence, risk factors, and molecular characteristics of MRSA and MSSA, specifically USA300 strain. | Trend towards higher MRSA overall in US-born patients (p=0.11), but not statistically significant.<br>US-born patients had a significantly higher prevalence of MRSA USA300 SSTIs.<br>**Racial disparities** in MRSA cases: higher proportion of Black US-born patients and Hispanic non-US-born patients. Risk factors for MRSA: male gender, sharing hygiene products, and animal exposure. Non-US-born patients had a higher prevalence of MSSA infections. |
| Immergluck et al. (2019) | USA (Atlanta) | 2002-2010 | Retrospective epidemiology study | Children treated for *Staphylococcus aureus* infections at two paediatric hospitals in the Atlanta MSA | Standardized incidence ratios (SIR) of CA-MRSA and CA-MSSA; odds ratios for CA-MRSA infection | **Racial disparities:** Black children had significantly higher odds of CA-MRSA (aOR 1.58, 95% CI 1.44-1.75, $p < 0.0001$).<br>**Age:** Children <4 years had increased CA-MRSA risk (aOR 1.65, 95% CI 1.48-1.83, $p < 0.0001$).<br>**Socioeconomic factors:** Higher CA-MRSA odds in children with no/public insurance ($p < 0.01$). Neighbourhood crowding ($p < 0.001$) and poverty (significant association) linked to CA-MRSA. |
| Casey et al (2012) | USA (Pennsylvania) | 2001-2010 | Retrospective cohort study | Patients with primary-care providers within the Geisinger Health System.<br>□ | Incidence of HA-MRSA and CA-MRSA<br>CA-MRSA.<br>Incidence of skin and soft tissue infections (SSTIs).<br>Risk factors for MRSA and SSTIs. | **Age:** Older age increased risk for HA-MRSA; younger age increased risk for CA-MRSA. Community Socioeconomic<br>**Deprivation:** Higher deprivation linked to increased odds of both HA-MRSA and CA-MRSA. |
| Blakiston & Freeman (2020) | Aotearoa New Zealand | N/A | Ecological study | 18 geographically distinct populations (District Health Boards) | Incidence rate of MRSA infection | **Ethnicity:** Māori & Pacific positive correlating with MRSA infections, while European ethnicity was negatively correlated.<br>Household crowding, socioeconomic **deprivation**, **age** <5 years was positively correlated with MRSA infections |
| Oates et al. (2020) | USA | 2016 | Cross-sectional observational study | Pediatric patients with cystic fibrosis | MRSA prevalence | Neighbourhood **deprivation:** Doubled the odds of MRSA [OR 2.26 (1.14–4.45), $P < 0.05$].<br>Sociodemographic: Deprivation correlated with rural residence, lack of parental college education, and low household income. |
| Kirby and Herbert (2013) | 15 European countries | 2003-2010 | Ecological study | Populations of 15 European countries | Antimicrobial resistance (EARS-Net), antimicrobial consumption (ESAC), income inequality (SWIID) | Strong positive correlation between income inequality and methicillin resistance ($r = 0.87$). |
| Restrepo et al. (2010) | USA (San Antonio, TX) | 1999-2002 | Retrospective cohort study | Hispanic and non-Hispanic White patients hospitalized with community-acquired pneumonia (CAP) | MRSA prevalence | No significant difference in MRSA rates between Hispanic and non-Hispanic White patients. |
| ***Streptococcus pneumoniae*** |  |  |  |  |  |  |
| Restrepo et al. (2010) | USA (San Antonio, TX) | 1999-2002 | Retrospective cohort study | Hispanic and non-Hispanic White patients hospitalized with community-acquired pneumonia (CAP) | MRSA prevalence, other drug-resistant pathogens, clinical outcomes (mortality, length of stay, ICU admission) | **Ethnicity:** Penicillin-resistant *S. pneumoniae* -Significantly higher rate in Hispanic patients (21.7%) compared to non-Hispanic White patients (0%). No significant difference in macrolide-resistant *S. pneumoniae* or overall drug-resistant *S. pneumoniae* between the two groups. |
| Kirby and Herbert (2013) | 15 European countries | 2003-2010 | Ecological study | Populations of 15 European countries | Resistance to penicillin and macrolides in *S. pneumoniae*, income inequality (SWIID) | **S. pneumoniae:** Weak positive correlation between income inequality and resistance to penicillin ($r = 0.34$). Weak positive correlation between income inequality and resistance to macrolides ($r = 0.28$). |
| ***Escherichia coli*** |  |  |  |  |  |  |
| Mitrani-Gold et al (2023) | USA | 2015-2019 | Retrospective cohort study | Female outpatients aged $\geq 12$ years with *E. coli* positive urine culture | 1)Fluoroquinolone-not-susceptible (FQNS) (*Intermediate/resistant E. coli* isolates. – 2)Multidrug-resistant (*MDR E. coli* isolates (NS to $\geq 1$ drug across $\geq 3$ classes). | 1)Age was found to be an independent risk factor for patients having FQNS *E. coli* isolates.<br>- Patients aged 25-50 years had a significantly higher likelihood of *FQ NS E. coli* (OR 2.7, CI 1.0-7.4), compared to patients aged 12-17 years.<br>-Patients older than 50 years had a significantly higher likelihood of *FQ NS E. coli* (OR 5.1, CI 1.9-14.3), compared to patients aged 12-17 years.<br>2) Age was not found to be an independent predictor of MDR *E. coli*. |
| Casey et al. (2021) | USA (California) | 2015-2017 | Case-control and case-case study | Patients with *E. coli* UTI | MDR *E. coli* UTI, Medicaid use, interpreter use, census tract deprivation | MDR prevalence: 10.4% (KPSC), 12.8% (Sutter Health). Interpreter use: Increased MDR risk (RR 1.36 KPSC, RR 1.28 Sutter). Medicaid use: Increased MDR risk (8% KPSC, 9% Sutter). High census tract deprivation: Increased MDR risk (RR 1.04 KPSC, RR 1.14 Sutter). Weak association between sociodemographic factors and overall UTI. |
| Robey et al. (2017) | UK | 2013-2014 | Retrospective study | Patients with *E. coli* positive cultures | Antibiotic susceptibility, ESBL production, age-specific trends | ESBL: Highest at extremes of age, decreases up to age 3, then increases. Amoxicillin, co-amoxiclav, ciprofloxacin: Resistance decreases initially, then increases with age. |
|  |  |  |  |  |  | Trimethoprim, trimethoprim-sulfamethoxazole: Resistance increases steadily, pronounced in adults >85 years.<br>Cephalexin: Resistance increases after age 55. |
| Kirby and Herbert (2013) | Europe | 2003-2010 | Ecological study | Populations of 15 European countries | Resistance to various antibiotics in *E. coli*, income inequality (SWIID) | **E. coli:** Moderate to strong positive correlations between income inequality and resistance to: Aminoglycosides (r = 0.84) Aminopenicillins (r = 0.73) . Third-generation cephalosporins (r = 0.70) Fluoroquinolones (r = 0.71). Weak positive correlation between income inequality and resistance to carbapenems (r=0.34). |
| Blakiston & Freeman (2020) | New Zealand |  | Ecological study | 18 geographically distinct populations defined by district health boards in Aotearoa New Zealand. | Incidence rate of ESBL-E. coli infection. | Positive correlations between the incidence rate of ESBL-E. coli infection and household crowding, community antimicrobial use, Asian ethnicity, Pacific ethnicity, and overseas-born new arrivals. A negative correlation was observed between the incidence rate of ESBL-E. coli infection and European ethnicity. |
| ***Helicobacter Pylori*** |  |  |  |  |  |  |
| Hsiang, J., et al (2013) | New Zealand (South Auckland) | February 2012 - October 2012 | Prospective, single-centre study | Patients undergoing gastroscopy in South Auckland | Prevalence of *H. pylori* infection, antibiotic resistance, eradication rate with standard triple therapy. | **Ethnicity:** High *H. pylori* prevalence in Māori (34.8%), Pacific People (31.3%), and Orientals (23.8%) compared to Europeans (7.7%) and Indians (19.2%). **Ethnicity-Linked Antibiotic Resistance:** Clarithromycin resistance (≥15%) predominantly found in Māori, Pacific People, and Orientals, directly impacting treatment efficacy.<br>**Ethnicity-Dependent Treatment Outcomes:** Lower eradication rates with OAC therapy in ethnic groups with high clarithromycin resistance (63.9%) versus low resistance groups (85.7%).<br>**Clarithromycin Resistance and Ethnicity:** Clarithromycin-resistant *H. pylori* strains showed only 33.3% eradication success, primarily found in specific ethnic groups.<br>**Overall Eradication Success:** 84.8%, but significantly influenced by ethnicity-related antibiotic resistance. |
| ***Other Pathogens (Enterococcus, Klebsiella, and Pseudomonas)*** |  |  |  |  |  |  |
| Kirby and Herbert (2013) | 15 European countries | 2003-2010 | Ecological study | Populations of 15 European countries | Resistance to various antibiotics, income inequality (SWIID) | **E. faecalis:** Moderate positive correlations between income inequality and resistance to: Aminopenicillins (r = 0.54) <Vancomycin (r = 0.73) High-level gentamicin (r = 0.62)<br>**E. faecium:** Moderate positive correlation between income inequality and resistance to: Vancomycin (r = 0.72) Weak<br>positive correlation between income inequality and resistance to: High-level gentamicin ($r = 0.38$),Aminopenicillins ($r = 0.26$)<br>**K. pneumoniae:** Moderate positive correlations between income inequality and resistance to: Aminoglycosides ($r = 0.50$), Third-generation cephalosporins ($r = 0.57$) Fluoroquinolones ($r = 0.50$). Weak positive correlation between income inequality and resistance to carbapenems ($r = 0.33$)<br>**P. aeruginosa:** Moderate positive correlations between income inequality and resistance to: Aminoglycosides ($r = 0.51$) Carbapenems ($r = 0.50$) Ceftazidime ($r = 0.51$) Fluoroquinolones ($r = 0.53$) . Weak positive correlation between income inequality and resistance to: Piperacillin-tazobactam ($r = 0.46$) Amikacin ($r = 0.21$) |

Most papers examined both the hospital and community settings. Among the 18 included studies, the most frequently identified pathogens were *Staphylococcus (13),* followed by *Escherichia* (5). Other species included *Streptococcus (2), Enterococci (1), Klebsiella (1), Pseudomonas (1) and Helicobacter (1)*.

Ethnicity was the most frequently cited element of health inequality followed by deprivation and age. Other factors included income level and migration status.

### Staphylococcus aureus

Studies on *S. aureus* were primarily conducted in the USA (n=9)^22,23,23,35,26.27.28.29,30^ with two from Europe^31,32^, and one each from Australia^33^ and New Zealand.^34^

Several U.S. studies highlighted racial disparities in MRSA infection rates, with Black patients experiencing significantly higher rates than White patients, even after adjusting for factors such as age, sex, race, and income.^22,23^

Gualandi *et al*. examined MRSA disparities in the USA and found that, despite a decline in hospital-associated (HA-MRSA) and hospital-associated community-onset (HACO) MRSA rates from 2005 to 2014, Black patients still had 2-3 times higher incidence rates. For chronic dialysis patients, although the racial disparity was reduced, it remained significant (adjusted rate ratio: 1.83, 95% CI: 1.72–1.96).^22^

Similarly, Ray *et al*. found increased MRSA infection risk among African-Americans and Hispanics compared to White patients. In contrast, Asian patients were less likely to develop MRSA. Disparities were also observed by age, with individuals over 65 at higher risk compared to those aged 18-49, and by socioeconomic status, with those in lower-income census blocks at increased risk.^23^

Iwamoto *et al*. conducted a trend analysis in children, showing that MRSA disproportionately affects young infants and Black children.^24^ Additionally, a prospective study in Australia showed that Indigenous Australians were 29.2 times more likely to have community-associated MRSA (CA-MRSA) compared to non-Indigenous Australians.^33^

See *et al.* conducted a retrospective cohort study in the USA to examine whether area-based socioeconomic factors might explain racial differences in community MRSA infection incidence. Findings showed that annual invasive community-associated MRSA incidence was 4.59 per 100,000 among White and 7.60 per 100,000 among Black population (RR, 1.66). However, after accounting for education, income, housing value and rural status, 91% of the original racial disparity was explained and no significant association of Black race with community-associated MRSA remained.^25^

Jenks *et al.* found that immigrants in New York City had lower, though not statistically significant, MRSA infection rates, but significantly higher MSSA wound infection rates, compared to USA-born citizens.^26^ Conversely, Immergluck *et al.* reported that USA-born patients were significantly more likely to have *S. aureus* skin and soft tissue infections (SSTIs) caused by the MRSA USA300 strain.^27^ They further revealed racial disparities within MRSA infections with a higher proportion of Black individuals among USA-born MRSA cases and a higher proportion of Hispanic individuals among non-USA-born MRSA cases.^27^

Auguet *et al*. conducted a spatial analysis in London to investigate the relationship between socioeconomic deprivation and CA-MRSA and HA-MRSA. The findings showed that both CA-MRSA and HA-MRSA were associated with household deprivation (CA-MRSA, Risk Ratio (RR) 1.72; HA-MRSA, RR 1.57). CA-MRSA was also associated with household overcrowding (RR 1.58) and broader socioeconomic barriers. Additionally, CA-MRSA was associated with recent immigration to the UK (RR 1.7) and residents living near deprived areas were at higher risk of infection and/or colonisation compared to those living near more affluent areas.^31^

Similarly, a spatial analysis study in the US found that CA-MRSA was more prevalent in areas with a higher proportion of younger children (<5 years), Black residents, and households experiencing crowding.^28^ Additionally, Casey *et al*. found that higher community socioeconomic deprivation was associated with increased odds of both HA-MRSA and CA-MRSA.^29^

An ecological study in New Zealand also found positive correlations between MRSA incidence and factors like household crowding, socioeconomic deprivation, Māori and Pacific ethnicity, and children under 5-years old.^34^

Oates *et al*. conducted a cross-sectional study which found that neighbourhood deprivation was a significant risk factor for MRSA, with paediatric cystic fibrosis patients in deprived areas being twice as likely to contract MRSA compared to those in more affluent areas.^30^ Similarly, a European study examining income inequality and drug-resistant infections found a strong association between income inequality and MRSA infection.^32^

### Streptococcal infections

Two Streptococcal studies were included in this review, one conducted in the USA^35^ and the other in Europe.^32^ In the USA, a retrospective cohort study of patients diagnosed with community associated pneumonia found higher rates of penicillin-resistant *S. pneumoniae* in Hispanic patients compared to non-Hispanic White patients. However, no significant differences were found between these two groups for other drug-resistant *S. pneumoniae* infections, 30-day mortality, ICU admission, or length of stay.^35^ In the European study examining the relationship between income inequality and AMR, a weak association was found between *Streptococcus pneumoniae* resistant to penicillin and macrolides and income inequality.^32^

### Escherichia infections

Five studies on *Escherichia* infections from the USA^36,37^, UK^37^, Europe^38^, and New Zealand^34^ were included.

In a case-control study conducted by Casey *et al*. in the USA, the impact of individual and community-level sociodemographic factors on the risk of multidrug-resistant (MDR) *E. coli* urinary tract infections (UTIs) was explored. The study found that Medicaid use (a proxy for low income) and the need for an interpreter (proxy for migrant status) were significantly associated with an increased risk of MDR *E. coli* UTIs. Additionally, living in areas with high census tract-level deprivation was linked to a higher risk of MDR *E. coli* UTIs.^36^

A retrospective study in the USA identified age as an independent risk factor for fluoroquinolone antibiotic non-susceptibility, with adults aged 25-50 years and over 50 years showing a significantly higher risk compared to adolescents aged 12-17. However, age was not found to be a risk factor for MDR *E.coli* infections.^37^ In the UK, a retrospective study reported strong positive associations between *E. coli* antibiotic non-susceptibility and older age adults (>85-years) and infants (<3-years). Resistance patterns varied by antibiotic: trimethoprim resistance steadily increased across all ages, while resistance to amoxicillin and co-amoxiclav decreased initially and later increased with age. Extended-spectrum beta-lactamase (ESBL)-producing *E. coli* was most prevalent in the youngest and oldest age groups.^38^

The European study showed that *E. coli* resistance to aminoglycosides was strongly associated with income inequality (r=0.84, 95% CI 0.80-0.88) whilst resistance to aminopenicillins, fluoroquinolones and third generation cephalosporins was moderately associated with income inequality (r=0.73, 95% CI 0.70-0.77, r=0.71 95% CI 0.6-0.82 and r=0.7, 95% CI 0.68-0.73 respectively). Resistance to carbapenems was weakly associated with income inequality (r=0.34, 95% CI 0.18-0.49).^32^

An ecological study in New Zealand found strong positive correlations between the incidence of ESBL-producing *E. coli* infections and crowded housing conditions, and moderate positive correlations with Asian and Pacific ethnicity, overseas-born status, and community antimicrobial use.^34^

### Helicobacter infection

One study on *H. pylori* infection in New Zealand was included.^39^ The study revealed that the prevalence of *H. pylori* infection varied by ethnicity, with the highest prevalence in Māori populations (34.9%), followed by Pacific People (29.6%), Orientals (23.8%), and Indians (9.3%). New Zealand European populations had the lowest prevalence (7.7%). Resistance to *H. pylori* treatment was found in metronidazole (49.3%), clarithromycin (16.4%), and moxifloxacin. No significant difference in metronidazole resistance was observed between ethnic groups or by birthplace; however, clarithromycin resistance was found in 25% of the New Zealand-born indigenous Māori population.^39^

### Other pathogens

The following pathogens (Enterococcus, Klebsiella, and Pseudomonas) were examined in the same study,^32^ which explored the relationship between income inequality and AMR across 15 European countries between 2003 and 2010.

### Enterococcus

Resistance of *E. faecilis* to aminopenicillins was moderately associated with income inequality (r = 0.54, 95% CI 0.49-0.60). Resistance to vancomycin (r = 0.73, 95% CI 0.68-0.79) and high-level gentamicin (r = 0.62, 95% CI 0.55-0.69) was also moderately associated with income inequality. Vancomycin resistance in *E. faecium* was moderately associated with income inequality (r = 0.72, 95% CI 0.64-0.80), while resistance to high-level gentamicin (r = 0.38, 95% CI 0.17-0.58) and aminopenicillins (r = 0.26, 95% CI 0.06-0.44) showed weak associations.

### Klebsiella

Resistance of *K. pneumoniae* to aminoglycosides, third-generation cephalosporins, and fluoroquinolones was moderately associated with income inequality (r = 0.50, 95% CI 0.42-0.57; r = 0.57, 95% CI 0.54-0.60; r = 0.50, 95% CI 0.42-0.57, respectively). Carbapenem resistance showed a weak association (r = 0.33, 95% CI 0.29-0.37).

### Pseudomonas infection

Resistance of *P. aeruginosa* to aminoglycosides, carbapenems, ceftazidime, and fluoroquinolones was moderately associated with income inequality (r = 0.51, 95% CI 0.47-0.56; r = 0.50, 95% CI 0.45-0.55; r = 0.51, 95% CI 0.47-0.55; r = 0.53, 95% CI 0.48-0.58). Resistance to piperacillin-tazobactam and amikacin was weakly associated (r = 0.46, 95% CI 0.37-0.55 and r = 0.21, 95% CI 0.15-0.26, respectively).

## Discussion

To the best of our knowledge, at the time the study was initiated, it was the first scoping review assessing the evidence on the association of health inequality factors (age, income, deprivation, ethnicity) and the risk of developing an antibiotic-resistant infection. We reviewed 18 studies published since 2010 in high-income countries, of which the majority of studies were conducted in the USA and focused on *Staphylococcus aureus*. Ethnicity was the most frequently assessed factor, followed by deprivation and age. Overall, the evidence showed that ethnic populations, those living in deprived areas and certain age groups are at a higher risk of an antibiotic-resistant infection.

### Ethnicity

The evidence from the US showed that Black, African-American and Hispanic populations were more likely to be infected with MRSA compared to White populations,^22,23^ whilst Asian populations were less likely to contract MRSA compared to White populations. This was the case, even after accounting for age, sex and other demographic factors. Interestingly, Gualandi *et al.*^22^ used chronic dialysis patients as a partial control, which reduced racial disparities but did not eliminate them, thus suggesting that racial disparities for HACO-MRSA may be attributed to differences in underlying health and access to care. See *et al.* showed that invasive MRSA infection rates were higher in Black compared to White populations, however, after accounting for socioeconomic factors there was no significant association of Black race with MRSA infection rates.^25^ Thus, highlighting the complex relationship between factors of health inequalities and AMR, and demonstrating the importance of considering a number of socioeconomic factors when explaining racial disparities. Data suggests that socioeconomic factors modify the relationship between race and MRSA infections and that systemic racism may be a mechanism that explains the association of Black race with MRSA infections.^40^ However, it is important to note that a wide range of socioeconomic data aren’t always readily available especially at the individual level. In New Zealand, Māori or pacific ethnicity was positively correlated with MRSA infection and ESBL *E. coli* infection.^34^ *H. pylori* infection varied by ethnicity and resistance to clarithromycin was seen in 25% of New Zealand born indigenous Māori populations.^39^

Only one study in the US assessed the association of ethnicity in relation to drug resistant Streptococcus pneumoniae infection and showed that Hispanic patients had higher rates of penicillin resistant pneumonia compared to non-Hispanic patients^35^. This is important as amoxicillin monotherapy is the standard of care for community-acquired bacterial pneumonia^41^, and a diagnostic test is not routinely used to confirm antimicrobial susceptibility, putting this sub-population of patients at risk of treatment failure. No differences were observed in macrolide resistant and other antibiotics,^34^ which differed to findings from Metley *et al.* (published outside of time frame of this review), who showed that Black patients in Philadelphia had higher rates of macrolide resistant *S. pneumoniae*.^42^

### Deprivation

The review found seven studies which assessed the impact of deprivation on MRSA^23,25,28,29,30,31,34^ and two on *E.coli*^34,36^ from the US, UK and New Zealand. In the USA, individuals living in lower income census blocks^23^ and in areas with high deprivation^36^ were at an increased risk of MRSA infection and MDR *E. coli* UTIs, whilst high neighbourhood deprivation was a risk factor for MRSA in paediatric populations with cystic fibrosis.^30^ In the UK, CA-MRSA and HA-MRSA were both associated with deprivation in London^31^, whilst in New Zealand, crowded housing conditions strongly correlated with increased rates of ESBL-producing *E. coli* infections.^34^ This corroborated with findings from the English surveillance programme for antibiotic utilisation and resistance (ESPAUR) report, which show a link between deprivation, infection and AMR. Rates of resistant bloodstream infections (BSIs) in England were 42.6% higher in the lowest compared to the highest Index of Multiple Deprivation quintile in 2023 (38.1 *vs*. 26.7 infections/100,000 population); (p<0.005). Data from the 2023-24 ESPAUR report also suggests that these inequalities have worsened overtime with a 13.2% increase in the gap in rates of resistant bloodstream infections between the highest and lowest IMD quintiles since 2019.^43^ The reasons behind this may be multifactorial, including considerations for overall health, health literacy and access to healthcare.^44^

### Income inequality

One study showed that as income inequality increases in European countries, so do the rates of AMR for a number of species of bacteria, including *Enterococcus faecalis*, *E. coli*, *K. pneumoniae* and *S. aureus*. The correlation between income inequality and AMR was strongest for *S. aureus* and *E. coli*. In addition, the authors also showed that income inequality was positively correlated with antimicrobial consumption in outpatients, which in turn was positively correlated with resistance for multiple species-resistance pairings.^32^ Income inequality has also been associated with life expectancy, stress, heart disease, depression among others and within infectious diseases, deaths due to COVID-19 were found to be higher in US states with higher income inequality.^45^ An additional study showed that income inequality was positively associated with 30-day mortality rates for COVID-19 in 84 countries.^46^

### Age

Seven studies examined age as a factor in the incidence of MRSA^23,24,27,29,34^ and E.*coli.*^37,38^ In the USA, both older adults (> 65 years) and infants (< 1-year) were more likely to have MRSA compared to other age groups. Older adults were at increased risk for HA-MRSA which may be a result of the higher rates of hospital admissions within this age group.^47^ CA-MRSA was found to be higher in areas with a greater proportion of young children (< 5 years) in both the USA and New Zealand, suggesting a potential link between high antibiotic prescribing rates in this age group and increased CA-MRSA prevalence.^48^

Age was also found to be an independent risk factor for fluoroquinolone non-susceptible *E. coli* in the USA, with older adults more likely to be infected than adolescents. A similar trend toward increased prevalence of MDR *E. coli* with increasing age was also observed, although it did not reach statistical significance — possibly due to a limited sample size.^37^ In the UK, both older adults and younger infants were more likely to be infected by ESBL *E.coli*.^38^

Older individuals are generally more susceptible to infections and tend to receive antibiotics more frequently, which may contribute to the development of AMR. In addition, lifetime antibiotic exposure increases with age and may explain the higher prevalence of resistant isolates in older age groups.^49^ Further research is needed to understand the potential effect of age on risk of developing resistant infections and how it may be impacted by other interconnected elements of health inequity.

### Strengths and limitations

This study was initiated in 2021 as part of national activities for the UK AMR action plan and following the initiation of a specific health inequalities workstream within UKHSA which aimed to improve our understanding of disparities in resistant infection and antibiotic use within population with factors associated with health inequalities. To the best of our knowledge no scoping or systematic reviews were available that focused on the pathogens being monitored within the NAP. While this review filled an important gap in the literature and highlighted research gaps, there are some limitations to consider. Firstly, as this is a scoping review a critical appraisal was not undertaken. Other methodological limitations are only including papers published in English and those published in peer-reviewed journals, therefore publication bias cannot be excluded. Papers may also have been omitted in error as only two databases were searched, however a snowball technique was utilised to find additional papers. Secondly, all the studies included were observational and therefore causality cannot be inferred. As outlined throughout, simple cause-effect relationships across health inequalities are rarely the case as they are complex and are characterised by multiple outcomes and interactions. Therefore, even though most of the included studies assessed for a few confounders, an intersectional relationship of how health inequalities may interact together wasn’t addressed. In addition, some studies assessed relationships at an area level and therefore we cannot assume these findings hold true at the individual level: the ecological fallacy. Furthermore, all studies used different outcome measures which makes comparisons difficult. For example, even though some ethnic minorities were found to have higher rates of penicillin-resistant *S. pneumoniae* infection, the 30-day mortality, ICU admission and hospital length of stay did not differ between White populations, thus highlighting the importance of including a few different outcome measures to ascertain racial differences. Lastly, most included studies were conducted in the USA and focused on MRSA, thus limiting the generalisability of the results to other countries and pathogens.

Further assessment is required to determine the association of health inequalities and other pathogens, as most of the current research is focused on MRSA. Further studies are also needed to better understand the role of specific sociodemographic factors, other risk factors and the modifiability of these factors, as well as whether community intervention strategies can mitigate the identified risks.

## Conclusion

In summary, the results of this scoping review highlight disparities in Staphylococcus infections, particularly MRSA, related to race, ethnicity, age and socioeconomic factors. Similar disparities were observed in Streptococcus infections, while *H. pylori* infection prevalence varied among different ethnic groups. At the time of the review, the associations between income inequality and AMR in *Enterococcus, Escherichia, Klebsiella* and *Pseudomonas* suggest the need for addressing social determinants of health in combating AMR. These findings underscore the importance of considering the broader social and contextual factors in understanding and addressing bacterial infections and AMR.

## Transparency Declarations

None to declare

## Data Availability

All data produced in the present work are contained in the manuscript

