## Supplementary File for "Association of factors linked to health inequalities and the risk of antibiotic-resistant infection in high-income countries: a systematic scoping review"

Supplementary Material 1. Search strategy

21 (equit* or inequit* or nglish ru* or disparit* or equality).ti,kw. (69579)

2 (ethnic* or race or racial* or racis*).ti,kw. (83285)

3 ((social* or socio-economic or socioeconomic or economic or structural or material) adj3 (advantage* or disadvantage* or exclude* or exclusion or include* or inclusion or status or position or gradient* or hierarch* or class* or determinant*)).ti,kw. (34628)

4 (health adj3 (gap* or gradient* or hierarch*)).ti,kw. (1047)

5 (sociodemographic* or socio-demographic* or income or wealth* or poverty or educational level or level of education or educational attainment or well educated or better educated or unemploy* or home owner* or tenure or affluen* or well off or better off or worse off).ti,kw. (55333)

6 protected characteristic*.ti,ab,kw. (16)

7 exp *ancestry group/ (68686)

8 exp *"ethnic or racial aspects"/ (48765)

9 ethnic group/ (71560)

10 religion/ (70449)

11 sexual orientation/ (6044)

12 disability/ (115167)

13 race/ (61919)

14 sex/ (36302)

15 vulnerable population/ (20299)

16 income group/ (404)

17 lowest income group/ (30866)

18 medically uninsured/ (6311)

19 exp *socioeconomics/ (74705)

20 “social determinants of health”/ (10498)

21 social class/ (31970)

22 health care disparity/ (17475)

23 health disparity/ (23220)

24 *high risk population/ (5214)

25 homeless person/ (1975)

26 or/1-25 (719900)

27 exp *infection/ (2174509)

28 infection*.ti,kw. (635238)

29 infectious disease*.ti,kw. (30995)

30 *infectious complication/ (1827)

31 or/27-30 (2319030)

32 attributable risk/ (7317)

33 behavioral risk factor surveillance system/ (3522)

34 infection risk/ (91722)

35 mortality risk/ (25421)

36 patient risk/ (8413)

37 population risk/ (9111)

38 recurrence risk/ (80584)

39 *risk factor/ (92455)

40 *prevalence/ (74447)

41 or/32-40 (379768)

42 developed country/ (34466)

43 high income country/ (4842)

44 ((high-income or developed) adj countr*).ti,kw. (2302)

45 Great Britain.ti,kw. (3045)

46 united kingdom.ti,kw. (15508)

47 England.ti,kw. (22677)

48 United States/ (1153149)

49 (United States or America or USA).ti,kw. (127456)

50 or/42-49 (1266767)

51 26 and 31 and 41 and 50 (812)

52 limit 51 to ( nglish language and yr=”2010 -Current”) (526)

53 exp Coronavirinae/ (46979)

54 exp Coronavirus infection/ (126681)

55 Coronavirus disease 2019/ (107016)

56 ((corona* or corono*) adj1 (virus* or viral* or virinae*)).ti,ab,kw. (2640)

57 (coronavirus* or  nglish rus* or coronavirinae* or CoV).ti,ab,kw. (81420)

58 (“2019-nCoV*” or 2019nCoV* or “19-nCoV*” or 19nCoV* or nCoV2019* or “nCoV-2019*” or nCoV19* or “nCoV-19*” or “COVID-19*” or COVID19* or “COVID-2019*” or COVID2019* or “CORVID-19*” or CORVID19* or “WN-CoV” or WNCoV or “HcoV-19*” or HcoV19* or “HcoV-2019*” or HcoV2019* or CoV or “2019 novel*” or Ncov* or “n-cov” or “SARS-CoV-2*” or “SARSCoV-2*” or “SARSCoV2*” or “SARS-CoV2*” or SARSCov19* or “SARS-Cov19*” or “SARSCov-19*” or “SARS-Cov-19*” or SARSCov2019* or “SARS-Cov2019*” or “SARSCov-2019*” or “SARS-Cov-2019*” or SARS2* or “SARS-2*” or SARScoronavirus2* or “SARS-coronavirus-2*” or “SARScoronavirus 2*” or “SARS coronavirus2*” or SARScoronovirus2* or “SARS-coronovirus-2*” or “SARScoronovirus 2*” or “SARS coronovirus2*” or covid).ti,ab,kw. (126927)

59 or/53-58 (159343)

60 52 not 59 (401)

61 limit 60 to (books or chapter or conference abstract) (74)

62 60 not 61 (327)

63 exp antibiotic agent/ (1542232)

64 antibiotic*.ti,ab,kw. (476303)

65 exp antiinfective agent/ (3711115)

66 antimicrobial*.ti,ab,kw. (242045)

67 or/63-66 (3852009)

68 exp prescription/ (215110)

69 prescribing.ti,ab,kw. (78121)

70 prescription*.ti,ab,kw. (179716)

71 regimen*.ti,ab,kw. (414240)

72 or/68-71 (709137)

73 26 and 50 and 67 and 72 (428)

74 73 not 59 (425)

75 limit 74 to ( nglish language and yr=”2010 -Current”) (313)

76 limit 75 to (books or chapter or conference abstract) (110)

77 75 not 76 (203)

78 exp antibiotic resistance/ (177712)

79 ((antibiotic or antimicrobial) adj resistan*).ti,ab,kw. (86760)

80 78 or 79 (202913)

81 31 or 41 (2599020)

82 26 and 50 and 80 and 81 (121)

83 82 not 59 (121)

84 limit 83 to ( nglish language and yr=”2010 -Current”) (78)

85 limit 84 to (books or chapter or conference abstract) (12)

86 84 not 85 (66)

Supplementary Material 2. Study Characteristics and source

| **Author** | **Country** | **Journal** | **Pathogen** | **Source of study** | **Health Inequality** |
|---|---|---|---|---|---|
| Ray et al (2012) | USA | Journal of clinical microbiology | *Staphylococcus Aureus* | First search | Ethnicity, Age, Deprivation |
| Gualandi et al (2018) | USA | Clinical infectious diseases: an official publication of the Infectious Diseases Society | *Staphylococcus Aureus* | First search | Ethnicity |
| Iwamoto et al (2013) | USA | Paediatrics | *Staphylococcus Aureus* | Snowballing | Ethnicity, Age |
| Tong et al. (2012) | Australia | BMC Infectious Diseases | *Staphylococcus Aureus* | Snowballing | Ethnicity |
| See et al (2017) | USA | Clinical Infectious Diseases | *Staphylococcus Aureus* | Snowballing | Deprivation, Ethnicity |
| Jenks (2016) | USA | Travel medicine and infectious disease | *Staphylococcus Aureus* | First search | Migrant populations |
| Auguet et al. (2016) | UK | PLOS Medicine | *Staphylococcus Aureus* | Snowballing | Deprivation |
| Immergluck et al. (2023) | USA | Antibiotics (Basel) | *Staphylococcus Aureus* | Second search | Migrant population, Ethnicity |
| Casey et al (2012) | USA | Epidemiology and Infection. | *Staphylococcus Aureus* | Snowballing | Deprivation, age |
| Immergluck et al. (2019) | USA | BMC Infectious Diseases | *Staphylococcus Aureus* | Snowballing | Age, Community and socioeconomic deprivation, Ethnicity |
| Oates et al. (2020) | USA | Paediatric Pulmonology | *Staphylococcus Aureus* | First search | Deprivation |
| Blakiston & Freeman (2020) | New Zealand | New Zealand Medical Journal | *Staphylococcus aureus*, *Escherichia coli* | Snowballing | Deprivation, Ethnicity, Age |
| Kirby and Herbert (2013) | Europe | PLoS One | *Staphylococcus aureus, Streptococcus pneumoniae, Escherichia coli, Enterococcus, Klebsiella, and Pseudomonas* | First search | Income inequality |
| Restrepo et al (2010) | USA | Hospital Practice | *Staphylococcus aureus, Streptococcus pneumoniae* | First search | Ethnicity |
| Casey et al (2021) | USA | Open Forum Infectious Diseases | *Escherichia coli* | Second search | Deprivation (census tract-level), Mediaid (proxy for income), language interpreter (proxy for migrant/vulnerable) |
| Mitrani-Gold et al (2023) | USA | PLOS One | *Escherichia coli* | Snowballing | Age |
| Robey et al (2017) | UK | Journal of Infection | *Escherichia coli* | Second search | Age |
| Hsiang, J. et al (2013) | New Zealand | New Zealand Medical Journal | *Helicobacter Pylori* | First search | Ethnicity |

Supplementary Material 3. Preferred Reporting Items for Systematic reviews and Meta-Analyses extension for Scoping Reviews (PRISMA-ScR) Checklist

| SECTION | ITEM | PRISMA-ScR CHECKLIST ITEM | REPORTED ON PAGE # |
|---|---|---|---|
| **TITLE** | | | |
| Title | 1 | Identify the report as a scoping review. | 1 |
| **ABSTRACT** | | | |
| Structured summary | 2 | Provide a structured summary that includes (as applicable): background, objectives, eligibility criteria, sources of evidence, charting methods, results, and conclusions that relate to the review questions and objectives. | 2-3 |
| **INTRODUCTION** | | | |
| Rationale | 3 | Describe the rationale for the review in the context of what is already known. Explain why the review questions/objectives lend themselves to a scoping review approach. | 3 - 4 |
| Objectives | 4 | Provide an explicit statement of the questions and objectives being addressed with reference to their key elements (e.g., population or participants, concepts, and context) or other relevant key elements used to conceptualize the review questions and/or objectives. | 4 |
| **METHODS** | | | |
| Protocol and registration | 5 | Indicate whether a review protocol exists; state if and where it can be accessed (e.g., a Web address); and if available, provide registration information, including the registration number. | NA |
| Eligibility criteria | 6 | Specify characteristics of the sources of evidence used as eligibility criteria (e.g., years considered, language, and publication status), and provide a rationale. | 5, 7 |
| Information sources* | 7 | Describe all information sources in the search (e.g., databases with dates of coverage and contact with authors to identify additional sources), as well as the date the most recent search was executed. | 5 |
| Search | 8 | Present the full electronic search strategy for at least 1 database, including any limits used, such that it could be repeated. | Supplementary material 1 |
| Selection of sources of evidence† | 9 | State the process for selecting sources of evidence (i.e., screening and eligibility) included in the scoping review. | 6 |
| Data charting process‡ | 10 | Describe the methods of charting data from the included sources of evidence (e.g., calibrated forms or forms that have been tested by the team before their use, and whether data charting was done independently or in duplicate) and any processes for obtaining and confirming data from investigators. | 6 |
| Data items | 11 | List and define all variables for which data were sought and any assumptions and simplifications made. | 6 |
| Critical appraisal of individual sources of evidence§ | 12 | If done, provide a rationale for conducting a critical appraisal of included sources of evidence; describe the methods used and how this information was used in any data synthesis (if appropriate). | NA |

| SECTION | ITEM | PRISMA-ScR CHECKLIST ITEM | REPORTED ON PAGE # |
|---|---|---|---|
| Synthesis of results | 13 | Describe the methods of handling and summarizing the data that were charted. | 6 |
| **RESULTS** | | | |
| Selection of sources of evidence | 14 | Give numbers of sources of evidence screened, assessed for eligibility, and included in the review, with reasons for exclusions at each stage, ideally using a flow diagram. | 8 |
| Characteristics of sources of evidence | 15 | For each source of evidence, present characteristics for which data were charted and provide the citations. | 10-15 |
| Critical appraisal within sources of evidence | 16 | If done, present data on critical appraisal of included sources of evidence (see item 12). | NA |
| Results of individual sources of evidence | 17 | For each included source of evidence, present the relevant data that were charted that relate to the review questions and objectives. | 16-20 |
| Synthesis of results | 18 | Summarize and/or present the charting results as they relate to the review questions and objectives. | 16-20 |
| **DISCUSSION** | | | |
| Summary of evidence | 19 | Summarize the main results (including an overview of concepts, themes, and types of evidence available), link to the review questions and objectives, and consider the relevance to key groups. | 20-23 |
| Limitations | 20 | Discuss the limitations of the scoping review process. | 23-24 |
| Conclusions | 21 | Provide a general interpretation of the results with respect to the review questions and objectives, as well as potential implications and/or next steps. | 25 |
| **FUNDING** | | | |
| Funding | 22 | Describe sources of funding for the included sources of evidence, as well as sources of funding for the scoping review. Describe the role of the funders of the scoping review. | 25 |

PRISMA-ScR = Preferred Reporting Items for Systematic reviews and Meta-Analyses extension for Scoping Reviews.
